# Breast and Cervical Cancer Screening Rates in Student-Run Free Clinics: A Systematic Review

**DOI:** 10.1101/2022.03.31.22273259

**Authors:** Sophia Ying Xiao, Catherine Kendall Major, Katie A. O’Connell, David Lee, Christine Lin, Esther Sarino, Kevin Chen

## Abstract

**Objective:** To assess rates of breast and cervical cancer screening at student-run free clinics to better understand challenges and strategies for advancing quality and accessibility of women’s health screening at student-run free clinics.

**Data sources:** We performed a systematic search of Ovid MEDLINE, PubMed, Web of Science, and Google Scholar databases from database inception to 2020 using keywords related to student-run clinics, breast cancer screening, and cervical cancer screening.

**Study eligibility criteria:** We included all English-language publications describing screening rates of breast and/or cervical cancer at student-run free clinics within the United States. Five authors screened abstracts and reviewed full texts for inclusion.

**Study appraisal and synthesis methods:** Two reviewers extracted data independently for each publication using a structured data extraction table. Disagreements were resolved by group consensus. Two reviewers then assessed for risk of bias for each text using a modified Agency for Healthcare Research and Quality checklist for cross-sectional and prevalence studies. Results were synthesized qualitatively due to study heterogeneity.

**Results:** Of 3634 references identified, 12 references met study inclusion criteria. The proportion of patients up-to-date on breast cancer screening per guidelines ranged from 45% to 94%. The proportion of patients up-to-date on cervical cancer screening per guidelines ranged from 40% to 88%.

**Conclusion:** Student-run free clinics can match breast and cervical cancer screening rates amongst uninsured populations nationally, though more work is required to bridge the gap in care that exists for the underinsured and uninsured.

## Introduction

A National Health Interview Survey (NHIS) from 2020 revealed that 64% of insured women were up-to-date for breast cancer screening compared to 30% of uninsured women.^1^ Similarly, a 2015 NHIS revealed that 82% of insured women were up-to-date with their cervical cancer screening compared to 78% of uninsured women.^2^ Both cancers can be detected by screening tests, with early detection and intervention reducing morbidity and mortality. Yet, screening rates among racial and ethnic minorities, low-income, uninsured, and underinsured women in the United States (US) remain low.^3,4^

One setting in which low-income, uninsured, and underinsured women may seek healthcare is in student-run free clinics (SRFCs), which are clinics led by students aiming to provide healthcare to underserved populations. For many of these patients, SRFCs may be their only source of care.^5,6^ As of 2014, SRFCs collectively provided over 37,000 annual patient visits. Quality of chronic disease management in SRFCs has been assessed, with evidence suggesting comparable care to other care settings.^5,7–15^ However, the efficacy of SFRCs at meeting national averages in women’s cancer screenings has shown mixed results. One study found greater rates of mammography among their patients compared to national averages.^16^ This same clinic also found higher rates of cervical cancer screening compared to national rates, while other clinics did not.^17,18^

## Objective

The objective of this systematic review was to examine rates of breast and cervical cancer screenings at SRFCs. From this, we aim to better understand challenges and strategies to effectively improve quality of women’s health screening at SRFCs.

## Methods

### Information sources and search strategies

We performed a systematic search of Ovid MEDLINE, PubMed (National Library of Medicine), Web of Science (Clarivate), and Google Scholar (Google) databases from database inception to 2020 using key terms including: student-run clinic; human papillomavirus; women’s health; gynecology; breast cancer; cervical cancer; mammography; Papanicolaou test; Pap smear; preventative medicine; screening. The search strategy (**Appendix 1**) was created in conjunction with a medical librarian.

### Eligibility criteria and study selection

We included English-language studies describing screening rates of breast cancer and/or cervical cancer in adult women at SRFCs in the United States as primary or secondary outcomes. We included original observational and interventional studies published as peer-reviewed journal articles, abstracts, and theses.

We excluded studies if they did not report on the proportion of eligible women for receiving screening.

Five authors independently screened study titles and abstracts for inclusion, with each piece being reviewed by two authors. Conflicts were resolved by group consensus. Then, five authors reviewed complete texts of potentially eligible pieces, again with each piece reviewed by two authors and conflicts resolved by discussion.

The reference list of each included text was also reviewed to identify potentially eligible pieces not identified in the database searches.

### Data extraction and synthesis

For each included study, two authors independently used a structured data extraction form to collect information regarding the study characteristics, findings, and limitations. Disagreements were resolved in the same manner as described for study inclusion.

We organized studies related to breast cancer screening and cervical cancer screening separately. If a study related to both types of preventive care, the relevant data were included under their respective categories.

Due to heterogeneity of study designs, we synthesized results descriptively.

### Assessment of risk of bias

Additionally, two authors independently assessed for risk of bias for each study using the Agency for Healthcare Research and Quality (AHRQ) methodology checklist for cross-sectional and prevalence studies.^19^ This tool was chosen because all studies incorporated were primarily of cross-sectional design. Risk of bias was determined by the number of items on the checklist fulfilled by each study; the fewer the items fulfilled, the higher the risk of bias. Final quality assessment was based on consensus, and disagreements were resolved in the same manner as discussed for study inclusion.

## Results

### Study selection

Our search identified 3634 references. Of these, 3614 were excluded as either duplicates or because they did not meet our inclusion criteria based on title and abstract (n=3125). For the remaining 20 studies, we undertook full-text review and eliminated 8 studies because they did not specify the proportion of eligible women who had screening and as a result, we were not able to obtain rate of screening information from those studies.^20–27^ This left 12 references which ultimately met our inclusion criteria and were synthesized in the systematic review. **Figure 1** details the study selection process. Studies referred to breast and cervical cancer screening guidelines from either the United States Preventive Services Task Force (USPSTF) or the American Cancer Society (ACS).

**Figure 1.**
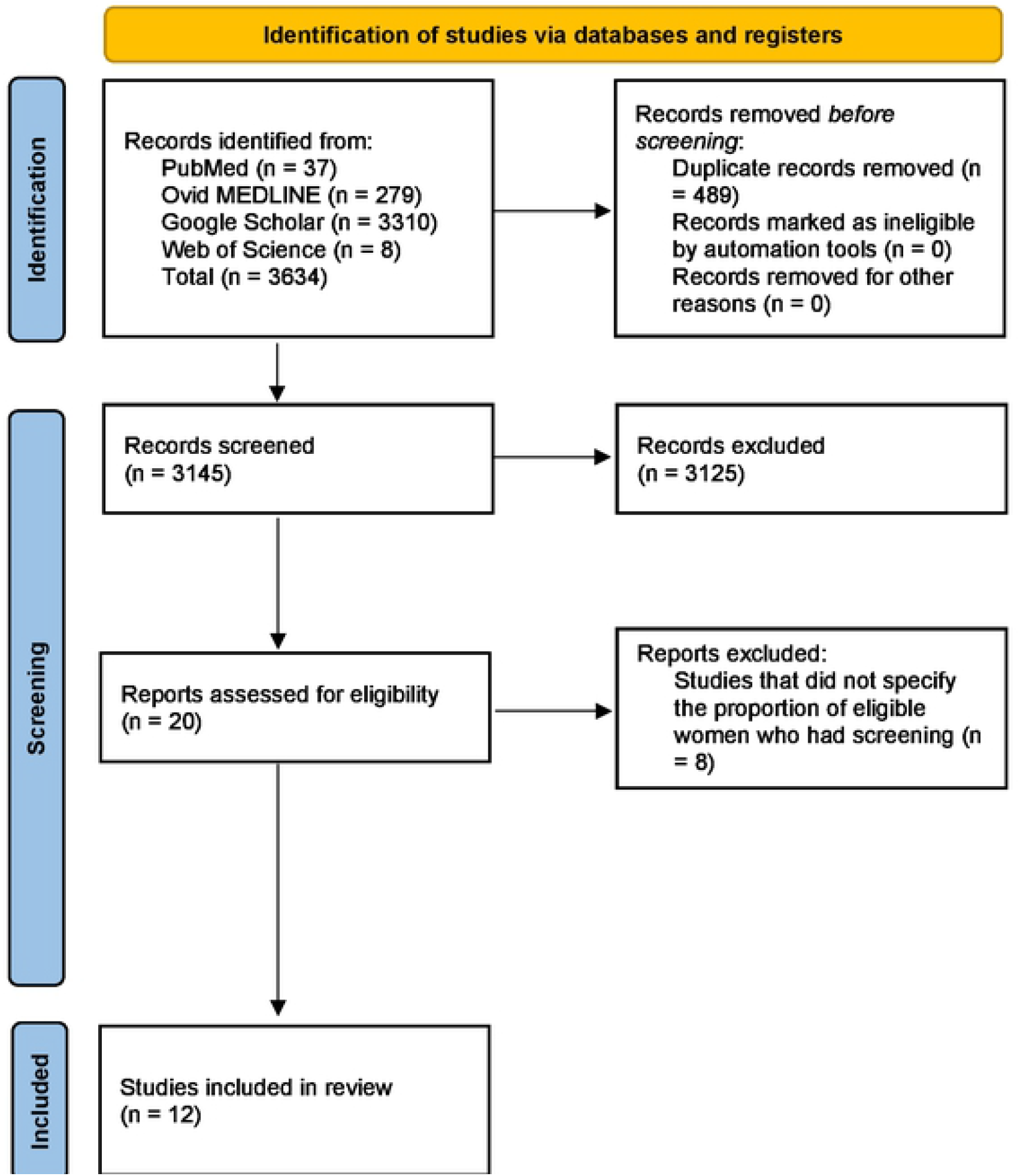
Flow diagram of study selection for inclusion in systematic review. Page MJ, McKenzie JE, Bossuyt PM, Boutron I, Hoffmann TC, Mulrow CD, et al. The PRISMA 2020 statement: an updated guideline for reporting systematic reviews. BMJ 2021;372:n71. doi: 10.1136/bmj.n71

### Breast cancer screening

#### Study characteristics

Eight studies reported on rates of breast cancer screening at SRFCs (**Table 1**). Breast cancer screening was performed via mammography for all studies. A total of 1408 patients were included in these studies. Two studies were cross-sectional questionnaires, two were cross-sectional chart reviews, and four were interventional studies.

**Table 1.**
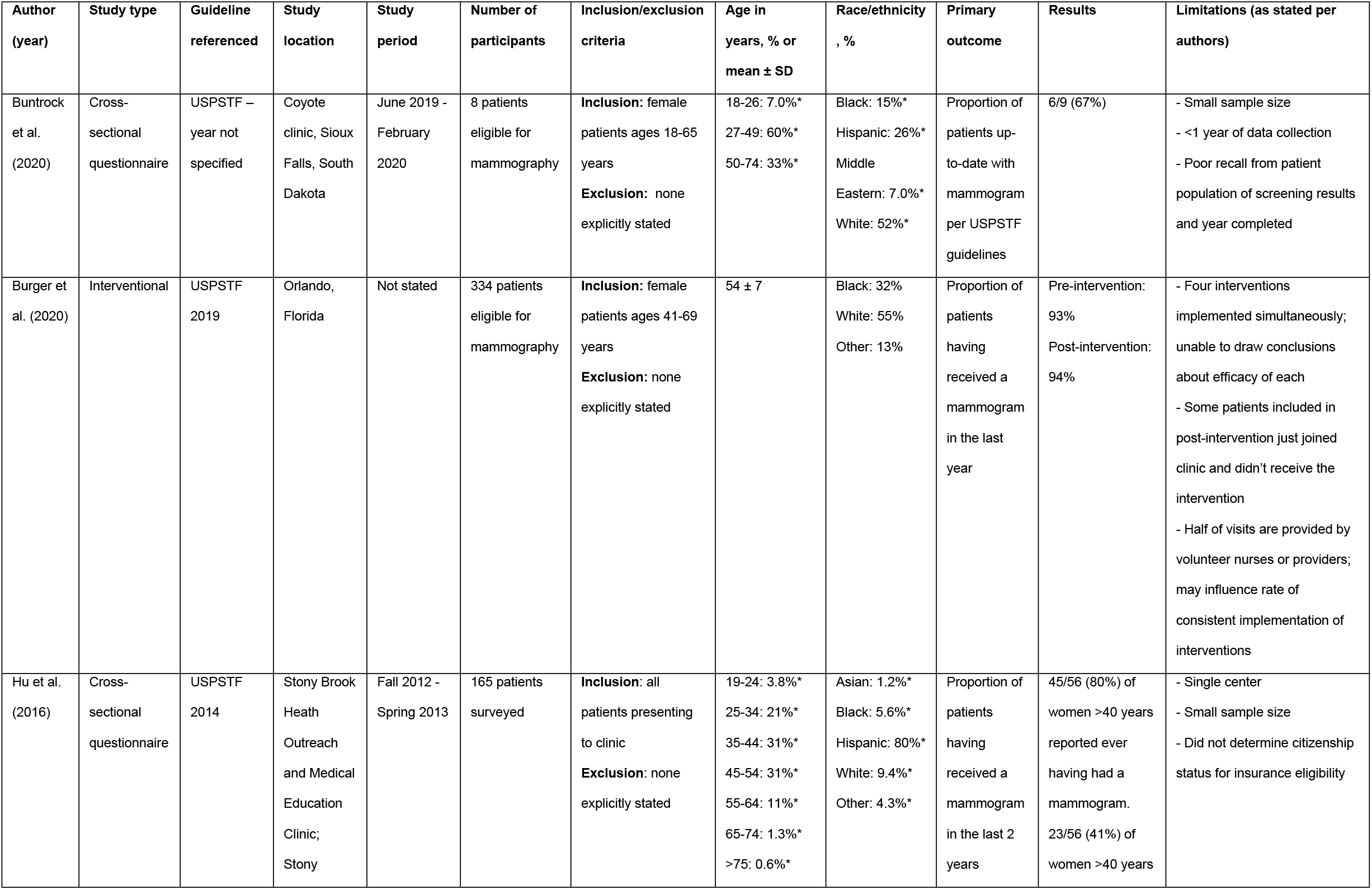
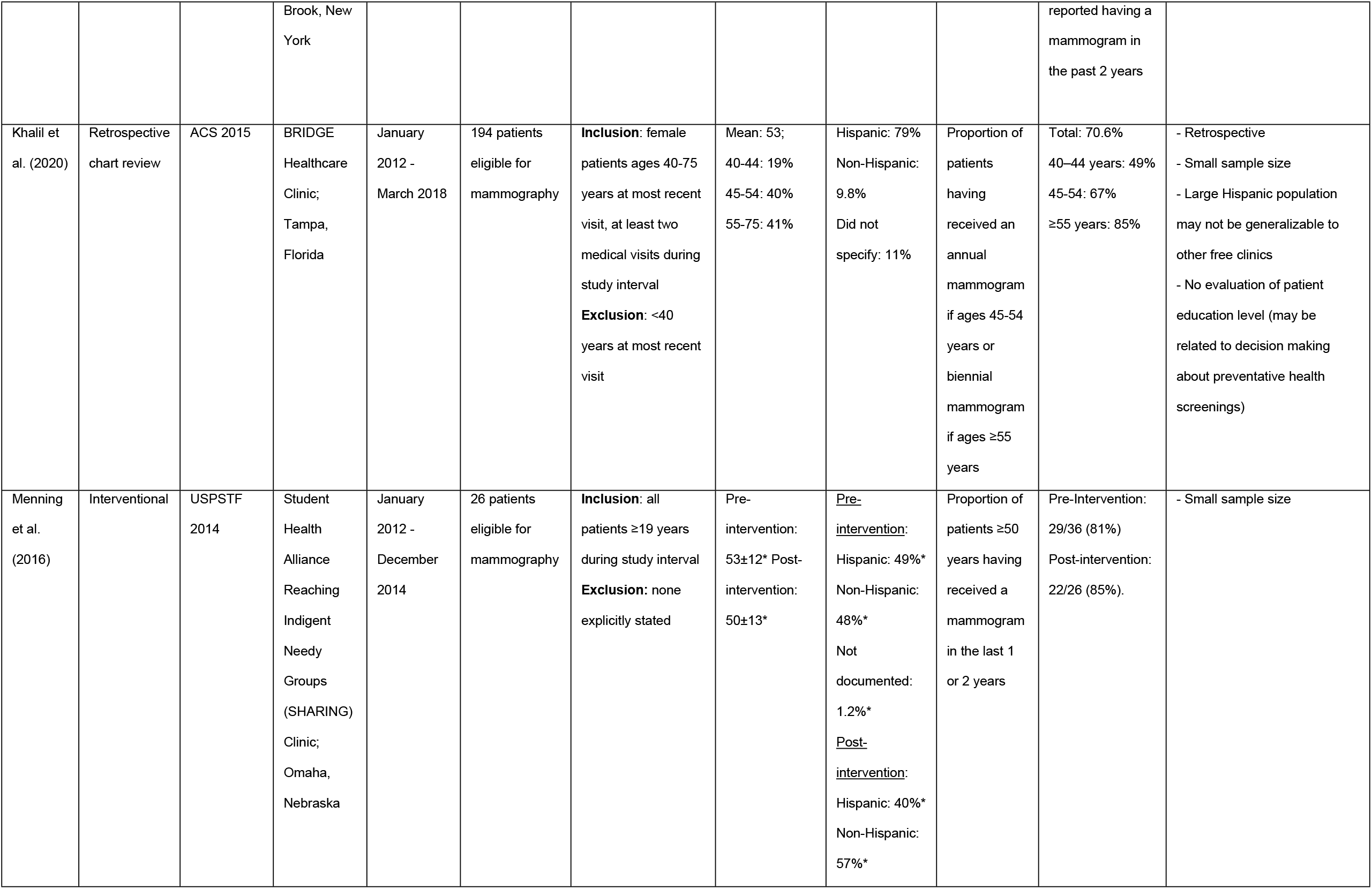
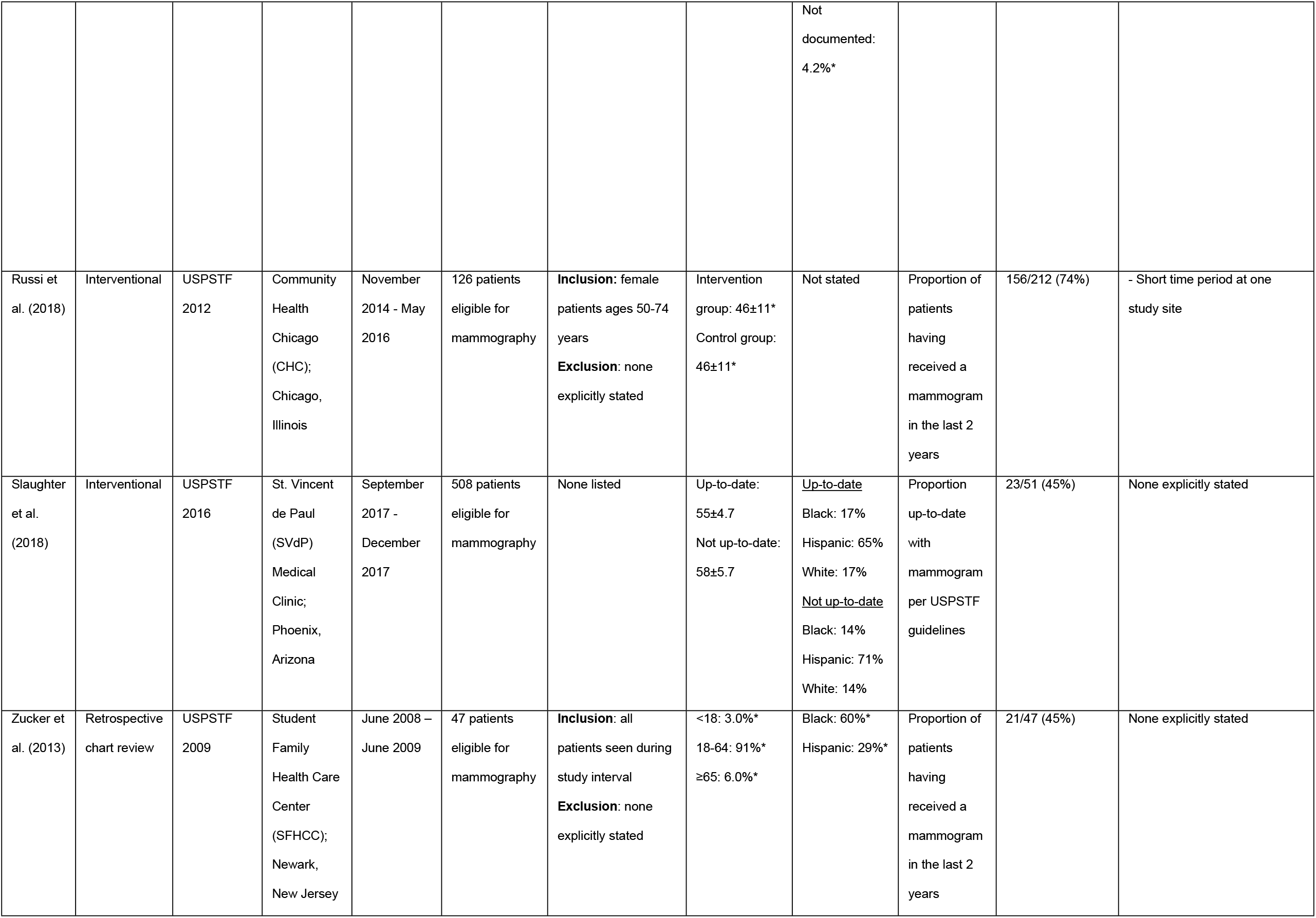
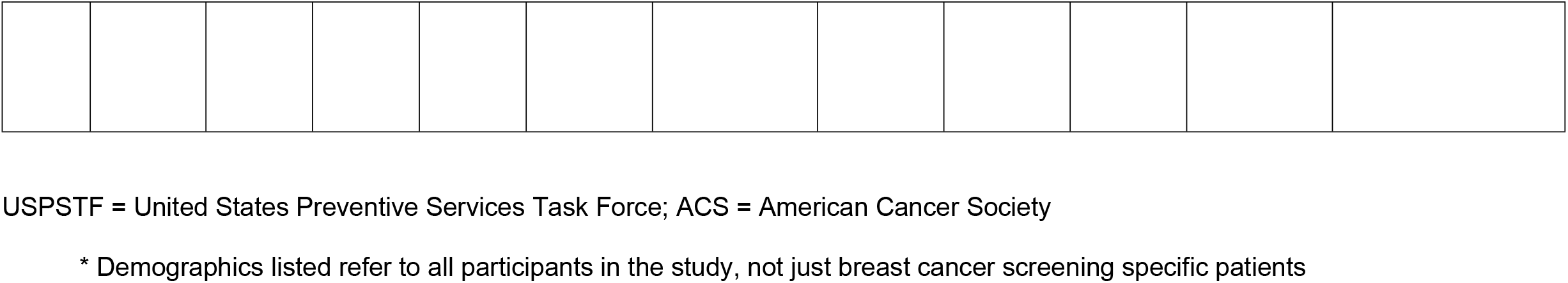
Details of studies that identified breast cancer screening rates in SRFCs via mammography

| Author (year) | Study type | Guideline referenced | Study location | Study period | Number of participants | Inclusion/exclusion criteria | Age in years, % or mean ± SD | Race/ethnicity, % | Primary outcome | Results | Limitations (as stated per authors) |
| --- | --- | --- | --- | --- | --- | --- | --- | --- | --- | --- | --- |
| Buntrock et al. (2020) | Cross-sectional questionnaire | USPSTF – year not specified | Coyote clinic, Sioux Falls, South Dakota | June 2019 - February 2020 | 8 patients eligible for mammography | **Inclusion:** female patients ages 18-65 years<br>**Exclusion:** none explicitly stated | 18-26: 7.0%*<br>27-49: 60%*<br>50-74: 33%* | Black: 15%*<br>Hispanic: 26%*<br>Middle Eastern: 7.0%*<br>White: 52%* | Proportion of patients up-to-date with mammogram per USPSTF guidelines | 6/9 (67%) | - Small sample size<br>- <1 year of data collection<br>- Poor recall from patient population of screening results and year completed |
| Burger et al. (2020) | Interventional | USPSTF 2019 | Orlando, Florida | Not stated | 334 patients eligible for mammography | **Inclusion:** female patients ages 41-69 years<br>**Exclusion:** none explicitly stated | 54 ± 7 | Black: 32%<br>White: 55%<br>Other: 13% | Proportion of patients having received a mammogram in the last year | Pre-intervention: 93%<br>Post-intervention: 94% | - Four interventions implemented simultaneously; unable to draw conclusions about efficacy of each<br>- Some patients included in post-intervention just joined clinic and didn't receive the intervention<br>- Half of visits are provided by volunteer nurses or providers; may influence rate of consistent implementation of interventions |
| Hu et al. (2016) | Cross-sectional questionnaire | USPSTF 2014 | Stony Brook Heath Outreach and Medical Education Clinic; Stony | Fall 2012 - Spring 2013 | 165 patients surveyed | **Inclusion:** all patients presenting to clinic<br>**Exclusion:** none explicitly stated | 19-24: 3.8%*<br>25-34: 21%*<br>35-44: 31%*<br>45-54: 31%*<br>55-64: 11%*<br>65-74: 1.3%*<br>>75: 0.6%* | Asian: 1.2%*<br>Black: 5.6%*<br>Hispanic: 80%*<br>White: 9.4%*<br>Other: 4.3%* | Proportion of patients having received a mammogram in the last 2 years | 45/56 (80%) of women >40 years reported ever having had a mammogram.<br>23/56 (41%) of women >40 years | - Single center<br>- Small sample size<br>- Did not determine citizenship status for insurance eligibility |
|  |  |  | Brook, New York |  |  |  |  |  |  | reported having a mammogram in the past 2 years |  |
| Khalil et al. (2020) | Retrospective chart review | ACS 2015 | BRIDGE Healthcare Clinic; Tampa, Florida | January 2012 - March 2018 | 194 patients eligible for mammography | **Inclusion:** female patients ages 40-75 years at most recent visit, at least two medical visits during study interval<br>**Exclusion:** <40 years at most recent visit | Mean: 53;<br>40-44: 19%<br>45-54: 40%<br>55-75: 41% | Hispanic: 79%<br>Non-Hispanic: 9.8%<br>Did not specify: 11% | Proportion of patients having received an annual mammogram if ages 45-54 years or biennial mammogram if ages ≥55 years | Total: 70.6%<br>40–44 years: 49%<br>45-54: 67%<br>≥55 years: 85% | - Retrospective<br>- Small sample size<br>- Large Hispanic population may not be generalizable to other free clinics<br>- No evaluation of patient education level (may be related to decision making about preventative health screenings) |
| Menning et al. (2016) | Interventional | USPSTF 2014 | Student Health Alliance Reaching Indigent Needy Groups (SHARING) Clinic; Omaha, Nebraska | January 2012 - December 2014 | 26 patients eligible for mammography | **Inclusion:** all patients ≥19 years during study interval<br>**Exclusion:** none explicitly stated | Pre-intervention: 53±12*<br>Post-intervention: 50±13* | <u>Pre-intervention:</u><br>Hispanic: 49%*<br>Non-Hispanic: 48%*<br>Not documented: 1.2%*<br><u>Post-intervention:</u><br>Hispanic: 40%*<br>Non-Hispanic: 57%* | Proportion of patients ≥50 years having received a mammogram in the last 1 or 2 years | Pre-Intervention: 29/36 (81%)<br>Post-intervention: 22/26 (85%). | - Small sample size |
|  |  |  |  |  |  |  |  | Not documented:<br>4.2%* |  |  |  |
| Russi et al. (2018) | Interventional | USPSTF 2012 | Community Health Chicago (CHC); Chicago, Illinois | November 2014 - May 2016 | 126 patients eligible for mammography | **Inclusion:** female patients ages 50-74 years<br>**Exclusion:** none explicitly stated | Intervention group: 46±11*<br>Control group: 46±11* | Not stated | Proportion of patients having received a mammogram in the last 2 years | 156/212 (74%) | - Short time period at one study site |
| Slaughter et al. (2018) | Interventional | USPSTF 2016 | St. Vincent de Paul (SVdP) Medical Clinic; Phoenix, Arizona | September 2017 - December 2017 | 508 patients eligible for mammography | None listed | Up-to-date: 55±4.7<br>Not up-to-date: 58±5.7 | <u>Up-to-date</u><br>Black: 17%<br>Hispanic: 65%<br>White: 17%<br><u>Not up-to-date</u><br>Black: 14%<br>Hispanic: 71%<br>White: 14% | Proportion up-to-date with mammogram per USPSTF guidelines | 23/51 (45%) | None explicitly stated |
| Zucker et al. (2013) | Retrospective chart review | USPSTF 2009 | Student Family Health Care Center (SFHCC); Newark, New Jersey | June 2008 – June 2009 | 47 patients eligible for mammography | **Inclusion:** all patients seen during study interval<br>**Exclusion:** none explicitly stated | <18: 3.0%*<br>18-64: 91%*<br>≥65: 6.0%* | Black: 60%*<br>Hispanic: 29%* | Proportion of patients having received a mammogram in the last 2 years | 21/47 (45%) | None explicitly stated |

USPSTF = United States Preventive Services Task Force; ACS = American Cancer Society
\* Demographics listed refer to all participants in the study, not just breast cancer screening specific patients

The percentage of White participants ranged from 9.4% to 55%. The percentage of Hispanic/Latino participants ranged from 26% to 79.5%. The percentage of Black participants ranged from 5.6% to 60%. One study had a 7% Middle-Eastern population.^28^ Another study had a 1.2% Asian population.^29^ For studies that listed participant ages specific to the sample of patients who were eligible for breast cancer screening, the majority fell between the ages of 40 and 50 years.^16,30,31^

Primary outcomes included a) date of last mammogram (n=1), b) rate of mammogram completion (n=5), c) percentage of patients up-to-date as per screening guidelines (n=2). Sample sizes ranged from 8-508 participants evaluated in each study.^31^

Of these eight studies, six (75%) used USPSTF guidelines to determine whether patients were up-to-date on their screenings, one used ACS guidelines, and one did not specify the guideline used.

#### Synthesis of results

For studies measuring the percentage of patients who are up-to-date on their screening as per guidelines, the proportion of patients who were screened appropriately amongst the studies ranged from 45% to 94%.

#### Risk of bias of included studies

The average number of checklist items fulfilled by each study reporting breast cancer screening data on the AHRQ methodology checklist was 4.0 (SD = 1.1) (**Table 2**). No studies explained patient exclusions from analysis or described how confounding was assessed or controlled. Only one study described assessments undertaken for quality assurance purposes.^17^ Only one study summarized patient response rates and completeness of data collection.^28^ Only two studies explain how missing data were handled in the analysis.^17,28^

**Table 2.** Risk of bias analysis

| *Author*<br>(Year) | *Define*<br>source of<br>information | *List inclusion and*<br>exclusion criteria<br>for exposed and<br>unexposed<br>subjects (cases<br>and controls) or<br>refer to previous<br>publications | *Indicate*<br>time<br>period<br>used for<br>identifying<br>patients | *Indicate*<br>whether<br>subjects were<br>consecutive if<br>not population-<br>based | *Indicate if*<br>evaluators of<br>subjective<br>components of<br>study were masked<br>to other aspects of<br>the status of the<br>participants | *Describe any*<br>assessments<br>undertaken for quality<br>assurance purposes<br>(e.g., test/retest of<br>primary outcome<br>measurements) | *Explain any*<br>patient<br>exclusions<br>from<br>analysis | *Describe*<br>how<br>confounding<br>was<br>assessed<br>and/or<br>controlled | *If applicable,*<br>explain how<br>missing data<br>was handled<br>in the<br>analysis | *Summarize*<br>patient response<br>rates and<br>completeness of<br>data collection | *Clarify what*<br>follow-up, if any,<br>was expected<br>and the<br>percentage of<br>patients for<br>which<br>incomplete data<br>or follow-up was<br>obtained |
| --- | --- | --- | --- | --- | --- | --- | --- | --- | --- | --- | --- |
| *Khalil et al.*<br>(2020) | + | + | + | + | NA | - | NA | - | NA | - | NA |
| *Menning et al.*<br>(2016) | + | + | + | + | NA | - | NA | - | NA | - | NA |
| *Price et al.*<br>(2019) | + | + | + | + | NA | - | NA | - | NA | - | NA |
| *Russi et al.*<br>(2018) | + | + | + | + | NA | - | NA | - | NA | - | NA |
| *Slaughter*<br>(2018) | + | - | + | + | NA | - | NA | - | NA | - | NA |
| *Buntrock et al.*<br>(2020) | + | + | + | + | NA | - | - | - | + | + | NA |
| *Burger et al.*<br>(2020) | + | + | - | + | NA | - | - | - | - | - | NA |
| *Butala et al.*<br>(2012) | + | + | + | + | NA | - | + | + | + | - | NA |
| *Butala et al.*<br>(2013) | + | + | + | + | NA | - | + | - | + | + | NA |
| *Hu et al.*<br>(2016) | + | - | + | + | NA | - | - | - | - | - | NA |
| *Veras et al.*<br>(2019) | + | - | + | + | NA | - | - | - | - | - | NA |
| *Zucker et al.*<br>(2013) | + | - | + | + | NA | + | - | - | + | - | NA |

+ = present; - = absent; N/A = not applicable

### Cervical Cancer Screening

#### Study characteristics

Ten studies reported on rates of cervical cancer screening at SRFCs (**Table 3**). Cervical cancer screening was performed via Papanicolaou (Pap) smear with cytology alone (without co-testing) for all studies. A total of 2198 patients were included in these studies. Two studies were cross-sectional questionnaires, four were cross-sectional chart reviews, and four were interventional studies. Of these ten studies, eight (80%) used USPSTF guidelines to determine whether or not patients were up-to-date on their screenings, one used ACS guidelines, and one did not specify the guideline used.

**Table 3.** Details of studies that identified cervical cancer screening rates in SRFCs via Pap smear.

| Author (year) | Study type | Guidelines referenced | Study location | Study period | # of participants | Inclusion/exclusion criteria | Age in years (mean ± SD) | Race/ethnicity | Primary outcome | Results | Limitations |
| --- | --- | --- | --- | --- | --- | --- | --- | --- | --- | --- | --- |
| Buntrock et al. (2020) | Cross-sectional questionnaire | USPSTF – year not specified | Coyote clinic, Sioux Falls, South Dakota | June 2019-February 2020 | 27 patients eligible for Pap smear | **Inclusion:** female patients ages 18-65 years<br>**Exclusion:** none explicitly stated | 18-26: 7.0%*<br>27-39: 60%*<br>50-74: 33%* | Black: 15%*<br>Middle Eastern: 7.0%*<br>Hispanic: 26%*<br>White: 52%* | Proportion of patients up-to-date with Pap smear per USPSTF guidelines | 21/27 (78%) | - Small sample size<br>- <1 year of data collection<br>- Poor recall from patient population of screening results and year completed. |
| Butala et al. (2012) | Retrospective chart review | USPSTF 2010-2011 | HAVEN Clinic; New Haven, Connecticut | October 2008-October 2009 | 55 patients eligible for Pap smear | **Inclusion:** female patients that kept at least one clinical visit during study period<br>**Exclusion:** no gender listed in patient chart, participants with missing patient charts (e.g., due to being under clinical use at the time of data abstraction) | 35 ± 10* | Black: 90%*<br>Latino: 90%*<br>Non-Latino White: 1.0%*<br>Other: 1.9%* | Proportion of patients up-to-date with Pap smear per USPSTF guidelines | 30/55 (55%) | - Missing charts<br>- Preventative services delivered at the clinic may not have been documented fully in the charts<br>- Nationwide comparisons may not be strictly comparable since BRFSS is a survey and this study was done via chart review<br>- HAVEN patients all have access to free care which may not be the case for all BRFSS patients. |
| Butala et al. (2013) | Interventional | USPSTF 2009 | HAVEN Clinic; New Haven, Connecticut | Pre-intervention: 2008-2009 | 166 patients eligible for Pap smear pre-intervention and 58 eligible | **Inclusion:** determined according to USPSTF guidelines on the | Pre-intervention: 36* | <u>Pre-intervention</u><br>Latino: 87%*<br>Other: 10%*<br><u>Post-intervention</u> | Proportion of patients up-to-date with Pap smear as | Pre-intervention: 68/116 (59%)<br>Post-intervention: 34/58 (59%) | - Observational pre/post design cannot establish a causal relationship between the implementation of the intervention and the increase |
|  |  |  |  | Post intervention: 2010-2011 | post-intervention | basis of age and gender<br>**Exclusion:** observations with missing age or gender; patients who were screening in a previous year within the guideline-recommended window | Post-intervention: 35* | Latino: 90%*<br>Other: 9.0%* | per USPSTF guidelines |  | in prevention, because data were combined from multiple years<br>- Patients may have repeated observations |
| **Hu et al. (2016)** | Cross-sectional questionnaire | Not stated explicitly | Stony Brook Health Outreach and Medical Education Clinic; Stony Brooke, New York | September 2012- May 2013 | 165 patients surveyed | **Inclusion:** all patients presenting to clinic<br>**Exclusion:** none explicitly stated | 19-24: 3.8%*<br>25-34: 21%*<br>35-44: 31%*<br>45-54: 31%*<br>55-64: 11%*<br>65-74: 1.3%*<br>>75: 0.6%* | Asian: 1.2%*<br>Black: 5.6%*<br>Hispanic: 80%*<br>White: 9.4%*<br>Other: 4.3%* | Proportion of patients having received a Pap smear in the past 3 years | 51/79 (65%) women >18 reported having a Pap smear in the past 3 years | - Single center<br>- Small sample<br>- No determined citizenship status |
| **Menning et al. (2016)** | Interventional | USPSTF 2014 | Student Health Alliance Reaching Indigent Needy Groups (SHARING | January 2012- December 2014 | 33 patients eligible for Pap smear | **Inclusion:** all patients ≥19 years during study interval<br>**Exclusion:** none explicitly stated | Pre-intervention: 53 ± 12*<br>Post-intervention: 50 ± 13* | <u>Pre-intervention:</u><br>Hispanic: 49%*<br>Non-Hispanic: 48%*<br>Not documented: 1.2%* | Proportion of patients up-to-date with Pap smear as per USPSTF guidelines | Pre-intervention: 25/38 (66%)<br>Post-intervention: 22/33 (67%) | - Small sample size |
|  |  |  | ) clinic;<br>Omaha,<br>Nebraska |  |  |  |  | <u>Post-</u><br><u>intervention:</u><br>Hispanic: 40%*<br>Non-Hispanic:<br>57%*<br>Not<br>documented:<br>4.2%* | pre-intervention<br>versus post-intervention |  |  |
| **Price et al.**<br>**(2020)** | Cross-sectional<br>chart review | ACS –<br>Cervical<br>2019 | Building<br>Relationshi<br>ps and<br>Initiatives<br>Dedicated<br>to Gaining<br>Equality<br>Healthcare<br>Clinic<br>(BRIDGE);<br>Tampa,<br>Florida | January<br>2012-March<br>2018 | 239 patients<br>eligible for Pap<br>smear | **Inclusion:** all female<br>patients aged 21 -<br>64 at their most<br>recent clinic visit with<br>2+ medical visits<br>during the study<br>interval.<br>**Exclusion:** history<br>of total hysterectomy | Mean 45;<br>21-29: 11%<br>30-64: 89% | Hispanic: 85%<br>Non-Hispanic:<br>7.1%<br>Did not specify:<br>7.9% | Proportion<br>of patients<br>up-to-date<br>with Pap<br>smear as<br>per ACS<br>guidelines | 210/239 (88%) | - Retrospective nature<br>- Small sample size<br>- Demographics not<br>necessarily generalizable |
| **Russi et al.**<br>**(2018)** | Intervention<br>al | USPSTF<br>2012 | Community<br>Health<br>Chicago<br>(CHC);<br>Chicago,<br>Illinois | November<br>2014-May<br>2016 | 212 patients<br>eligible for Pap<br>smear | **Inclusion:** for breast<br>cancer screening,<br>female patients<br>between 50 - 74, for<br>cervical cancer<br>screening, female<br>patients between 30<br>– 64. **Exclusion:**<br>none explicitly stated | Intervention<br>group: 46 ±<br>11*<br>Control group:<br>46 ± 11* | Not stated | Proportion<br>of patients<br>up-to-date<br>with Pap<br>smear as<br>per<br>USPSTF<br>guidelines | 156/212 (74%) | - Short time period at one<br>study site. |
| **Slaughter et al. (2018)** | Interventional | USPSTF cervical 2012 | St. Vincent de Paul (SVdP) Medical Clinic; Phoenix, Arizona | September 2017-December 2017 | 1164 patients eligible for Pap smear | None explicitly stated | Up to date: 43 ± 9.6<br>Not up to date: 51 ± 10 | <u>Up-to-date</u><br>Black: 5.4%<br>Hispanic: 88%<br>White: 6.9%<br><u>Not up-to-date</u><br>Black: 5.2%<br>Hispanic: 83%<br>White: 12% | Proportion of patients up-to-date with Pap smear as per USPSTF guidelines | 130/188 (69%) | None explicitly stated |
| **Veras et al. (2019)** | Retrospective chart review | USPSTF (no year stated) | Clinica Esperanza/ Hope Clinic (CEHC); Providence, Rhode Island | 2015-2017 | 75 patients eligible for Pap smear | **Inclusion:** patients presenting to clinic for non-gynecologic visits.<br>**Exclusion:** none explicitly stated | 21-65 | Hispanic (no % listed) | Proportion of patients up-to-date with Pap smear as per USPSTF guidelines | 40-62% | None explicitly stated |
| **Zucker et al. (2013)** | Retrospective chart review | USPSTF cervical 2003 | Student Family Health Care Center (SFHCC); Newark, New Jersey | June 2008-June 2009 | 62 patients eligible for Pap smear | **Inclusion:** all patients seen during study interval<br>**Exclusion:** none explicitly stated | < 18: 3.0%*<br>18-64: 91%*<br>≥ 65 y: 6.0%* | Black: 60%*<br>Hispanic: 29%* | Proportion of patients having received a Pap smear in the last 2 years | 32/62 (52%) | None explicitly stated |

USPSTF = United States Preventive Services Task Force; ACS = American Cancer Society
\* Demographics listed refer to all participants in the study, not just cervical cancer screening specific patients

The percentage of White participants ranged from 1.0% to 52%. The percentage of Hispanic/Latino participants ranged from 26% to 90%. The percentage of Black participants ranged from 5.2% to 89.5%. One study had participants with 7% Middle-Eastern population, and another reported a 1.2% Asian population.^28,29^ For studies that listed participant ages specific to the sample of patients who were eligible for cervical cancer screening, the majority fell between the ages of 30 and 50 years.^31–33^

Primary outcomes included a) date of last Pap smear (n = 1), b) Pap smear rate (n=3), c) percentage of patients up-to-date as per screening guidelines (n=6). Sample sizes ranged from 27-1164 participants evaluated in each study.^31^

#### Synthesis of results

For studies measuring the percentage of patients who are up-to-date on their screening as per guidelines, the proportion of patients who had been screened appropriately amongst the studies ranged from 40% to 88%.

#### Risk of bias of included studies

The average number of checklist items fulfilled by each study reporting cervical cancer screening data on the AHRQ methodology checklist was 4.6 (SD = 1.5) (**Table 2**). Only one of the 10 studies discussed assessments undertaken for quality assurance purposes.^17^ Only one study described how confounding was assessed or control.^18^ Only two studies summarized patient response rates and completeness of data collection.^28,34^ Only two studies explained patient exclusions from analysis.^21,24^

## Discussion

### Main findings

In this systematic review of breast and cervical cancer screening at SRFCs, we found that: (1) SRFCs largely use mammography and Pap smears with cytology alone for screening; (2) SRFCs may vary in the guidelines followed for screening interval recommendations but largely reference USPSTF and ACS; (3) adherence to guideline-recommended screening for breast and cervical cancer at SRFCs range from 45-94% and 40-88%, respectively, and; (4) most studies on this subject are at high risk of bias.

### Comparison with existing literature

In 2018, the national average of all eligible persons up-to-date with screening was 72.4% (95% CI 70.8–73.9) for breast cancer and 82.9% (95% CI 81.6–84.0) for cervical cancer.^35^ Within the uninsured population in the US, screening rates were 39.5% (95% CI 32.8–46.5) for breast cancer and 65.0% (95% CI 60.6–69.1) for cervical cancer.^35^ In our review, we found that approximately half of the studies included on breast cancer screening and one in ten of studies included on cervical cancer screening reported screening rates at or above the overall national average.^16,29,30,32,36^ If comparing with national screening rates for the uninsured only, all studies on breast cancer screening and five in ten studies on cervical cancer screening met or surpassed the national average.^28,31,32,36,37^ This suggests that, while there is substantial variation, people receiving care at SRFCs can achieve similar rates of breast and cervical cancer screening when compared with insured and uninsured populations receiving care at established settings, but further work is needed to close gaps in preventative care.

There may be several contributing factors to the difference between adherence to breast and cervical cancer screening recommendations at SRFCs. SRFCs rely on mostly volunteer faculty to oversee trainees, and not all faculty may have the same level of comfort with performing Pap smears versus referring patients for mammography. Another potential barrier to adherence to cervical cancer screening guidelines is ensuring that each clinic session has supplies to administer Pap smears and a pathology service available to interpret them. With breast cancer screening, where women are referred to an imaging facility that has a radiologist already available to interpret results, this may be less of an obstacle. Similarly, patients at SRFCs may not be as comfortable with trainees performing an invasive examination or test. The screening intervals for cervical cancer are also more infrequent than that for breast cancer screening, thus patients who have limited access to regular care may be more likely to be captured as adherent to cervical cancer screening than breast cancer screening if they receive intermittent screening for both. Other barriers to screening at SRFCs are broader. SRFCs usually need to partner with different diagnostic facilities to process laboratory and radiographic tests, and these tests may be of limited availability and cost. Furthermore, for patients who may have had intermittent access to care via SRFCs, addressing acute medical concerns or chronic disease management may displace focus on preventive services.

### Limitations

The studies synthesized in this review had several limitations. Many studies had small sample sizes. Most studies only fulfilled approximately 3 of 11 items on the AHRQ methodology checklist, suggesting a high risk of bias. Few studies summarized patient response rates and completeness of data collection or discussed assessments undertaken for quality assurance purposes or control/assessment of confounding. In addition, few studies provided demographic information specific to patients being evaluated for breast or cervical cancer screening rates. This may make it difficult to draw conclusions between national averages and SRFC averages. Future studies can benefit from increasing study size, discussing confounding, and improving discussion of patient demographics.

Our systematic review also has its limitations. Because SRFCs are organized by individual institutions in response to local community needs and student and supervising clinician interests, SRFCs tend to be heterogeneous in terms of scope of services, frequency of operation, and consistency of services provided. As a result, SRFCs often serve a diverse patient demographic with varying availability of services provided and less predictability of resources. Thus, findings from this systematic review may not be generalizable of SRFCs. Similarly, due to heterogeneity in study design, we were unable to perform a meta-analysis of breast and cervical cancer screening rates. In addition, some relevant studies may have eluded our search due to research remaining unpublished, being published in uncatalogued sources, or publication bias.

### Conclusions and implications

SRFCs are an important point of access for preventive care for the underserved. Our review suggests that SRFCs can match the rate of breast and cervical cancer screening in populations nationally. However, there is still a clear discrepancy between the rate of SRFC screening compared to the national insured population. Additional work is needed to capture data from SRFCs and improve screening in this setting.

## Data Availability

Data is extracted from public access articles published through Ovid Medline, Web of Science, and Google Scholar.

## Acknowledgements

No acknowledgements to disclose.

